# Longitudinal and socio-demographic trends in Long COVID and associated activity limitations: Results from the New York City Community Health Survey 2021 – 2023

**DOI:** 10.1101/2025.08.20.25334102

**Authors:** Samuel E. Packard, Amanda Lans, Yang Shi, Shadi Chamany

## Abstract

**Introduction:** In this study we describe the longitudinal and socio-demographic trends of Long COVID among adults in New York City using data from a large, population-based annual health survey.

**Methods:** Data from the 2021-2023 Community Health Survey included self-report long-term health effects of COVID-19. Survey-weighted prevalence of lifetime Long COVID (2021-2023), current Long COVID (2022-2023), and current Long COVID with activity limitations (2022-2023) were estimated by year and socio-demographic characteristics among the general adult population and among adults with prior COVID-19. Statistical significance was assessed at the p < 0.05 level.

**Results:** The prevalence of lifetime Long COVID increased in the general population during the study period (5.9% -14.3%, p < 0.001) but decreased as a share of adults with prior COVID-19 (34.6% - 26.3%, p < 0.001). In 2023 the population prevalence of current Long COVID was 11.6% and prevalence of related activity limitations was 7.6%, an increase from prior years affecting an estimated 501,000 - 766,000 New Yorkers. Inequities were observed by race/ethnicity, gender, education, and household poverty, and were more pronounced among adults with confirmed or suspected prior COVID-19.

**Discussion:** By 2023, 1 in 7 adults in New York City had experienced long-term health effects of COVID-19, half of whom reported current activity limitations. The prevalence and impact of Long COVID present an ongoing and important public health concern, especially given the disproportionate burden among people of color, cisgender women, transgender adults, and individuals with lower socio-economic status.

## Introduction

In response to early reports of persistent symptoms and incomplete recovery following acute COVID-19, now broadly referred to as Long COVID [1], the New York City Department of Health and Mental Hygiene (NYC Health Department) added questions about long-term physical and mental health effects of COVID-19 to its annual Community Health Survey (CHS) starting with the 2021 cycle. Surveillance of Long COVID is critical to understand and address the unequal impact of the pandemic, as the disabling nature of Long COVID may concentrate additional social and economic hardships in already marginalized communities [2]. We use data from the CHS to describe trends in the prevalence of Long COVID and associated activity limitations in New York City from 2021 – 2023.

## Methods

### Population and Study Design

The CHS is an annual cross-sectional survey of approximately 10,000 adults from an address-based sampling of NYC households [3]. Participants complete the survey online or by phone and data are weighted to the adult residential population. Data from the 2021-2023 CHS were used for this analysis, with missing responses for prior COVID-19 or Long COVID excluded via listwise deletion (0.5% of survey responses). This study was designated exempt human subjects research by the New York City Health Department Institutional Review Board.

### Definitions and Measures

Lifetime Long COVID was defined by responding affirmatively to the question “Have you experienced any long-term [physical health/emotional or mental health] effects that you think might be related to having had COVID-19?”, current Long COVID was defined as endorsing still feeling health effects at the time of the survey, and activity limitations was defined as responding “A little” or “A lot” to the question “How much difficulty do you have engaging in daily activities (or household responsibilities) because of these physical, mental, or emotional effects of COVID-19?” (Appendix Table 1). Lifetime Long COVID was collected in all three survey years, while current Long COVID and activity limitations were collected in 2022 and 2023.

### Statistical Analysis

Univariate analysis for each measure by year was followed by bivariate analysis of socio-demographic characteristics using pooled data from 2022 – 2023. Estimates were produced separately for the general adult population and adults with prior COVID-19 and were weighted to the New York City adult residential population based on the American Community Survey.

Between-year and between-group differences were assessed by a linear contrast t-test at the p < 0.05 level of significance. All analyses were conducted using SAS-callable SUDAAN version 11.0.4.

## Results

A total of 30,438 survey responses were included in the analysis. As shown in Table 1, the prevalence of lifetime Long COVID in the general adult population increased from 5.9% to 10.5% in 2021-2022. A subsequent increase in the prevalence of lifetime Long COVID (10.5% – 12.6% p = 0.008) as well as current Long COVID (9.6 – 11.6%, p = 0.007) and activity limitations (5.9% - 7.6%, p = 0.003) was observed in 2022-2023, and the proportion of adults who responded “Don’t Know” to both questions about long-term effects of COVID-19 increased from 1.4% to 4.9% during the study period. As a proportion of adults with prior COVID-19, the prevalence of lifetime Long COVID (32.8% - 26.3%, p < 0.001) and current Long COVID (26.9% – 21.4%, p < 0.001) decreased in 2022-2023.

**Table 1.** Prevalence of long-term health effects of COVID-19 (Long COVID) by year: NYC adults, 2021-2023.

| A) All adults | 2021 |  | 2022 |  | 2023 |  | 2021-2022 | 2022-2023 |
| --- | --- | --- | --- | --- | --- | --- | --- | --- |
|  | Weighted N | % (95% CI) | Weighted N | % (95% CI) | Weighted N | % (95% CI) | Δ % | Δ % |
| Lifetime Long COVID: Physical/Mental health | 377,000 | 5.9 (5.2-6.7) | 791,000 | 11.8 (10.7-12.9) | 943,000 | 14.3 (13.2-15.5) | ***5.9 | **2.5 |
| Lifetime Long COVID: Physical health | 329,000 | 5.1 (4.5-5.9) | 707,000 | 10.5 (9.6-11.6) | 826,000 | 12.6 (11.5-13.7) | ***5.4 | **2.0 |
| Lifetime Long COVID: Mental health | 195,000 | 3 (2.5-3.7) | 360,000 | 5.4 (4.7-6.2) | 455,000 | 6.9 (6.1-7.8) | ***2.3 | *1.5 |
| Lifetime Long COVID: Don't Know | 89,000 | 1.4 (1.0-1.9) | 181,000 | 2.7 (2.3-3.2) | 322,000 | 4.9 (4.2-5.7) | ***1.3 | ***2.2 |
| Current Long COVID |  |  | 647,000 | 9.6 (8.7-10.7) | 766,000 | 11.6 (10.6-12.7) |  | **2.0 |
| Current Long COVID Activity Limitations |  |  | 393,000 | 5.9 (5.1-6.7) | 501,000 | 7.6 (6.8-8.5) |  | **1.7 |
| B) Adults with prior COVID-19 | 2021 |  | 2022 |  | 2023 |  | 2021-2022 | 2022-2023 |
|  | Weighted N | % (95% CI) | Weighted N | % (95% CI) | Weighted N | % (95% CI) | Δ % | Δ % |
| Lifetime Long COVID: Physical/Mental health | 377,000 | 34.6 (31.1-38.4) | 791,000 | 32.8 (30.3-35.5) | 943,000 | 26.3 (24.4-28.3) | -1.8 | ***-6.5 |
| Lifetime Long COVID: Physical health | 329,000 | 30.3 (26.9-33.9) | 707,000 | 29.4 (26.9-32.0) | 826,000 | 23.2 (21.4-25.2) | -0.9 | ***-6.2 |
| Lifetime Long COVID: Mental health | 195,000 | 17.9 (15.1-21.2) | 360,000 | 14.9 (13.1-17.0) | 455,000 | 12.7 (11.2-14.3) | -3.0 | -2.2 |
| Lifetime Long COVID: Don't Know | 89,000 | 8.2 (6.1-10.9) | 181,000 | 7.5 (6.4-8.9) | 322,000 | 9.0 (7.8-10.4) | 0.7 | 1.5 |
| Current Long COVID |  |  | 647,000 | 26.9 (24.5-29.4) | 766,000 | 21.4 (19.6-23.2) |  | ** -5.5 |
| Current Long COVID Activity Limitations |  |  | 393,000 | 16.3 (14.4-18.4) | 501,000 | 14 (12.5-15.6) |  | -2.3 |
| Notes: Data are from the NYC Community Health Survey 2022-2023. Data are weighted to the adult residential population per the American Community Survey. |  |  |  |  |  |  |  |  |
| Confidence Intervals are a measure of estimate precision: the wider the CI, the more imprecise the estimate. P-values are shown as * < 0.05, ** < 0.01, and *** < 0.001. Weighted N population estimates are rounded to the nearest 1,000. |  |  |  |  |  |  |  |  |

The prevalence of Long COVID by socio-demographic characteristics is shown in Table 2. As a proportion of all adults, prevalence of lifetime and current Long COVID were higher among Latino adults (compared to white adults), cisgender women and transgender or non-binary adults (compared to cisgender men), adults with some college (compared to high school graduates), and was lower among older adults and high income households. Differences in the prevalence of activity limitations were similarly observed by race/ethnicity, gender, and household poverty, but not level of education or age group. As a proportion of adults with prior COVID-19, the prevalence of lifetime Long COVID, current Long COVID, and activity limitations were higher among all racial and ethnic groups compared to non-Latino white adults, higher among cisgender women and transgender or non-binary adults compared to cisgender men, lower among college graduates, and lower among higher income households.

**Table 2.** Prevalence of Long-term Health Effects of COVID-19 (Long COVID) by sociodemographic characteristics: NYC Adults, 2022 - 2023

|  | All adults<br>(Weighted N = 6,649,000) |  |  | Adults with prior COVID-19<br>(Weighted N = 3,004,000) |  |  |
| --- | --- | --- | --- | --- | --- | --- |
|  | Lifetime Long<br>COVID | Current Long<br>COVID | Current Long<br>COVID<br>Activity<br>Limitations | Lifetime Long<br>COVID | Current Long<br>COVID | Current Long<br>COVID<br>Activity<br>Limitations |
|  | % (95% CI) | % (95% CI) | % (95% CI) | % (95% CI) | % (95% CI) | % (95% CI) |
| Age Group |  |  |  |  |  |  |
| 18-24 | 12.4 (9.8-15.5) | *8.8 (6.7-11.4) | *5.1 (3.6-7.2) | 27.5 (22.2-33.5) | 19.5 (15.1-24.8) | 11.3 (8.0-15.7) |
| 25-29 | 14.4 (12.2-17.1) | 11.7 (9.7-14.1) | 6.6 (5.1-8.5) | 26.4 (22.5-30.7) | 21.4 (17.9-25.4) | 12.1 (9.4-15.4) |
| 30-44 (ref) | 14.5 (13.2-16.0) | 11.9 (10.7-13.3) | 7.3 (6.3-8.4) | 28.1 (25.7-30.7) | 23.0 (20.8-25.5) | 14.1 (12.2-16.1) |
| 45-64 | 14.2 (12.8-15.6) | 12.0 (10.8-13.4) | 7.7 (6.7-8.9) | *32.9 (30.1-35.9) | **27.9 (25.3-30.7) | *17.9 (15.7-20.4) |
| 65+ | ***9.6 (8.1-11.3) | ***8.3 (6.8-10.0) | 6.3 (5.0-7.8) | 28.0 (24.1-32.4) | 24.2 (20.4-28.5) | *18.4 (14.9-22.5) |
| Race/ethnicity |  |  |  |  |  |  |
| Asian/Pacific Islander | 13.1 (11.2-15.2) | 11.0 (9.3-13.0) | 7.4 (6.0-9.1) | **29.1 (25.3-33.2) | **24.4 (20.9-28.4) | **16.4 (13.4-20.0) |
| Black, non-Latino | 11.2 (9.6-13.0) | 9.6 (8.1-11.3) | 6.5 (5.2-7.9) | ***34.2 (29.8-38.8) | ***29.3 (25.2-33.7) | ***19.7 (16.2-23.8) |
| Latino | 15.6 (14.0-17.3) | ***13.1 (11.6-14.7) | ***8.5 (7.3-9.8) | ***36.5 (33.2-39.9) | ***30.6 (27.5-33.9) | ***19.8 (17.2-22.8) |
| Other, non-Latino | 14.5 (10.9-19.0) | 11.2 (8.3-14.9) | 6.7 (4.6-9.7) | **35.0 (27.3-43.6) | *27.1 (20.5-34.9) | *16.4 (11.3-23.1) |
| White, non-Latino (ref) | 12.4 (11.2-13.6) | 9.7 (8.7-10.9) | 5.7 (4.9-6.6) | 22.2 (20.2-24.3) | 17.4 (15.6-19.4) | 10.1 (8.7-11.7) |
| Gender |  |  |  |  |  |  |
| Cisgender Men (ref) | 10.2 (9.2-11.4) | 8.3 (7.3-9.3) | 5.1 (4.3-5.9) | 22.9 (20.7-25.2) | 18.5 (16.4-20.7) | 11.3 (9.7-13.2) |
| Cisgender Women | ***15.5 (14.5-16.6) | ***12.9 (11.9-13.9) | ***8.3 (7.5-9.2) | ***34.1 (32.1-36.2) | ***28.4 (26.5-30.4) | ***18.3 (16.6-20.0) |
| Transgender, Non-binary, or Other | **21.6 (15.2-29.7) | **17.8 (12.0-25.6) | **13.2 (8.7-19.4) | **45.0 (33.2-57.4)<sup>1</sup> | **37.2 (26.0-49.9)<sup>1</sup> | **27.6 (18.5-39.0) |
| Education |  |  |  |  |  |  |
| Less Than HS | 13.0 (10.9-15.4) | 10.9 (9.0-13.2) | 8.2 (6.5-10.2) | 39.7 (34.1-45.5) | 33.3 (28.1-39.0) | *25.0 (20.2-30.4) |
| HS or Equivalent (ref) | 12.2 (10.6-14.0) | 10.3 (8.8-11.9) | 6.6 (5.4-8.0) | 33.6 (29.7-37.7) | 28.3 (24.7-32.3) | 18.1 (15.0-21.7) |
| Some College | *15.2 (13.5-17.1) | *12.6 (11.0-14.3) | 8.0 (6.8-9.4) | 35.1 (31.6-38.8) | 29.0 (25.7-32.4) | 18.5 (15.8-21.5) |
| College Graduate | 12.8 (11.8-13.9) | 10.3 (9.4-11.3) | 6.0 (5.3-6.8) | ***22.5 (20.8-24.3) | ***18.1 (16.5-19.8) | ***10.5 (9.3-11.9) |
| Household Poverty |  |  |  |  |  |  |
| <100% | 14.2 (12.6-15.9) | 12.5 (11.0-14.2) | 8.5 (7.3-9.9) | 40.8 (37.0-44.8) | **36.0 (32.3-40.0) | 24.5 (21.2-28.1) |
| 100%-<200% (ref) | 13.6 (11.9-15.5) | 11.0 (9.5-12.8) | 7.8 (6.5-9.4) | 35.1 (31.2-39.3) | 28.6 (25.0-32.5) | 20.3 (17.1-23.9) |
| 200%-<400% | 13.8 (12.1-15.8) | 11.5 (9.9-13.3) | 7.0 (5.8-8.6) | 30.1 (26.6-33.9) | 25.1 (21.9-28.6) | *15.3 (12.7-18.4) |
| 400%-<600% | 13.5 (11.7-15.4) | 11.0 (9.4-12.8) | *5.9 (4.8-7.2) | ***25.3 (22.2-28.7) | **20.7 (17.8-23.9) | ***11.1 (9.0-13.5) |
| >= 600% | 11.0 (9.6-12.4) | **8.0 (6.9-9.4) | ***4.6 (3.7-5.7) | ***18.7 (16.5-21.1) | ***13.7 (11.8-15.9) | ***7.8 (6.3-9.6) |
| Notes: Data are from the NYC Community Health Survey 2022-2023. Data are weighted to the adult residential population per the American Community Survey. Confidence Intervals are a measure of estimate precision: the wider the CI, the more imprecise the estimate. P-values are shown as * < 0.05, ** < 0.01, and *** < 0.001. Weighted N population estimates are rounded to the nearest 1,000. |  |  |  |  |  |  |
| <sup>1</sup>Estimate should be interpreted with caution. Estimate's Relative Standard Error (a measure of estimate precision) is greater than 30%, or the 95% Confidence Interval half-width is greater than 10 or the sample size is too small, making the estimate potentially unreliable. |  |  |  |  |  |  |

## Discussion

Nearly 1 million adult New Yorkers in 2023 had ever experienced long-term health effects they thought were due to having had COVID-19, half of whom were still experiencing impacts to their day-to-day life. The magnitude of those affected and continue to have health problems disruptive to daily life makes this a substantial and ongoing public health concern.

The slowing increase in prevalence in the general population may be due to the evolution of SARS-CoV-2 variants, increased antibody seroprevalence, and lack of COVID-19 testing. The decrease in Long COVID as a share of adults with prior COVID-19 suggests that infections after 2022 were less likely to result in Long COVID, even as cumulative risk increases with reinfections [4].

While the incidence of Long COVID may be decreasing, its prevalence remains sustained and inequitably distributed. Socio-demographic inequities in Long COVID prevalence likely reflect social and structural determinants of health that inequitably influence risk factors and susceptibility, including underlying health conditions, limited access to medical care, and lack of paid sick leave. When restricting the analysis to those who self-reported COVID-19, socio- demographic inequities were more pronounced. This may be partly explained by lower rates of self-reported COVID-19 among people of color and those with lower socio-economic status (Appendix Table 2), which may indicate detection bias due to barriers to testing. These inequities should also be interpreted in the context of COVID-19 mortality patterns, as groups over-represented in COVID-19 deaths may be under-represented in estimates of Long COVID. Nevertheless, our findings show that people of color, women, transgender adults, and individuals with lower socio-economic status were more likely to experience long-term health outcomes after COVID-19, underscoring how structural inequities contribute to a disproportionate burden of both acute and long-term COVID-19 outcomes in marginalized communities.

### Limitations

Our findings are subject to several limitations. The case definition relies on subjective interpretation of long-term health effects related to COVID-19. Differential awareness of COVID-19’s long-term health impacts could bias estimates of differences over time or between groups. The case definition is also unique to this survey, limiting direct comparison with other studies [5]. Finally, while the survey design produces generalizable estimates for adult NYC residents, results may not be transferable to other populations.

## Conclusion

Local surveillance data are critical to promote awareness about the burden from Long COVID and support the continued need for public health measures such as vaccination, improved ventilation and filtration, and the use of high-quality masks to prevent new COVID-19 infections and reduce the incidence of Long COVID. The prevalence and impact of Long COVID underscores the importance of advocating for equal and fair access to resources for prevention, treatment, and protections such as workplace accommodations. This is especially important for communities that had less access to consistent and quality resources for prevention and treatment during the pandemic. Tailored, culturally appropriate interventions and coordination among healthcare providers, policymakers, and public health agencies are essential to mitigating the public health burden of Long COVID. The growing proportion of people uncertain about whether COVID-19 had long-term impacts on their health highlights the need for education for both the public and healthcare providers.

## Data Availability

Data availability and requests for public use data sets can be referenced at https://www.nyc.gov/site/doh/data/data-sets/community-health-survey-public-use-data.page

## Acknowledgements

We thank Sungwoo Lim, Gretchen van Wye, Toni Eyssallenne, and representatives from Patient-Led Research Collaborative for comments on a draft of this manuscript.

## Notes

**Funding:** This work was supported by a CDC Strengthening US Public Health Infrastructure, Workforce, and Data Systems grant (#NE11OE000057) and Epidemiology and Laboratory Capacity for Prevention and Control of Emerging Infectious Diseases (ELC) cooperative agreement (#NU50CK000517)

**Conflicts of Interest:** The authors report no conflicts of interest.

### Competing Interest Statement

The authors have declared no competing interest.

### Funding Statement

This work was supported by a CDC Strengthening US Public Health Infrastructure, Workforce, and Data Systems grant (#NE11OE000057) and Epidemiology and Laboratory Capacity for Prevention and Control of Emerging Infectious Diseases (ELC) cooperative agreement (#NU50CK000517)

### Author Declarations

This study was designated exempt human subjects research by the New York City Health Department Institutional Review Board (Protocol #24-088).

