## Supplemental Tables for "Longitudinal and socio-demographic trends in Long COVID and associated activity limitations: Results from the New York City Community Health Survey 2021 – 2023"

### Appendix

Appendix Table A-1: Survey questions and Long COVID measures

| Measure | Description | Response Options | Notes |
| --- | --- | --- | --- |
| Survey Questions | | | |
| Prior COVID-19 | “From February 2020 until now, do you think you may have had COVID-19?” | Yes  No  Don’t Know | Asked of all respondents |
| Ever Physical Health Effects | “Have you experienced any long-term physical health effects that you think might be related to having had COVID-19?” | Yes  No  Don’t Know | Asked if responded "Yes" to prior COVID-19 |
| Current Physical Health Effects | “Please tell us how much you agree or disagree with the following statement: I am still feeling long-term physical health effects of COVID-19” | Strongly Disagree  Somewhat Disagree  Neither Agree nor Disagree  Somewhat Agree  Strongly Agree | Asked if responded "Yes" to long-term physical health effects |
| Ever Mental Health Effects | “Have you experienced any long-term emotional or mental health effects that you think might be related to having had COVID-19?” | Yes  No  Don’t Know | Asked if responded "Yes" to prior COVID-19 |
| Current Physical Health Effects | “Please tell us how much you agree or disagree with the following statement: I am still feeling long-term emotional mental health effects of COVID-19” | Strongly Disagree  Somewhat Disagree  Neither Agree nor Disagree  Somewhat Agree  Strongly Agree | Asked if responded "Yes" to long-term mental health effects |
| Activity Limitations | “How much difficulty do you have engaging in daily activities (or household responsibilities) because of these physical, mental, or emotional effects of COVID-19?” | No Difficulty  A little Difficulty  A Lot of Difficulty | Asked if responded "Yes" to either long-term physical or mental health effects |
| Measures of Long COVID Prevalence | | | |
| Prevalence of lifetime Long COVID | Responded “Yes” to either Ever Physical Health Effects or Ever Mental Health Effects | Yes/No | Summary measure |
| Prevalence of current Long COVID | Responded “Strongly Agree” or “Somewhat Agree” to either Current Physical Health Effects or Current Mental Health Effects | Yes/No | Summary measure |
| Prevalence of current Long COVID Activity Limitations | Responded “A little difficulty” or “A lot of difficulty” to Activity Limitations and Current Physical/Mental Health Effects | Yes/No | Summary measure |

Appendix Table A-2: Prevalence of Prior COVID-19 by socio-demographic characteristics: NYC Adults, 2022 - 2023

|  | 2021 | | 2022 | | 2023 | |
| --- | --- | --- | --- | --- | --- | --- |
|  | Yes | Don't Know | Yes | Don't Know | Yes | Don't Know |
|  | % (95% CI) | % (95% CI) | % (95% CI) | % (95% CI) | % (95% CI) | % (95% CI) |
| Overall | 17.0 (15.8-18.3) | 12.2 (11.2-13.4) | 35.9 (34.4-37.5) | 14.4 (13.3-15.6) | 54.7 (52.9-56.4) | 9.8 (8.8-10.9) |
| Age Group |  |  |  |  |  |  |
| 18-24 | 21.6 (16.9-27.1) | 16.8 (12.5-22.2) | 38.9 (33.1-45.0) | 17.3 (13.0-22.6) | 49.0 (42.9-55.2) | 12.6 (9.3-16.8) |
| 25-29 | 20.6 (16.6-25.4) | 14.9 (11.6-19.0) | 46.5 (41.3-51.7) | 19.3 (15.5-23.8) | 63.8 (58.4-69.0) | 11.1 (7.9-15.4) |
| 30-44 (reference) | 17.5 (15.5-19.8) | 15.4 (13.2-18.0) | 43.2 (40.3-46.1) | 14.0 (12.2-15.9) | 60.5 (57.3-63.6) | 10.4 (8.7-12.4) |
| 45-64 | 18.5 (16.4-20.8) | 10.6 (9.0-12.6) | 31.4 (28.9-33.9) | 15.3 (13.3-17.6) | 54.6 (51.4-57.7) | 10.1 (8.4-12.3) |
| 65+ | 10.1 (8.2-12.3) | 5.2 (4.0-6.7) | 25.1 (22.0-28.5) | 9.0 (7.3-11.1) | 44.7 (41.0-48.6) | 6.3 (4.8-8.2) |
| Race/ethnicity |  |  |  |  |  |  |
| Asian/Pacific Islander | 11.2 (8.8-14.1) | 11.1 (8.8-14.0) | 31.4 (28.0-35.0) | 16.3 (13.6-19.3) | 59.0 (54.9-63.1) | 13.9 (11.1-17.1) |
| Black, non-Latino | 11.8 (9.5-14.6) | 12.2 (9.7-15.3) | 26.7 (23.2-30.5) | 12.8 (10.4-15.6) | 38.4 (34.6-42.4) | 9.3 (7.1-12.0) |
| Latino | 23.5 (20.9-26.3) | 13.5 (11.3-16.0) | 35.9 (32.7-39.2) | 13.8 (11.6-16.3) | 48.4 (44.9-52.0) | 10.4 (8.5-12.7) |
| Other, non-Latino | 15.5 (10.3-22.5) | 12.1 (7.6-18.8) | 32.1 (25.1-40.0) | 20.3 (14.6-27.6) | 50.9 (42.8-58.8) | 10.2 (6.0-16.9) |
| White, non-Latino (reference) | 17.8 (15.9-19.8) | 11.7 (10.2-13.6) | 44.1 (41.7-46.6) | 14.5 (12.8-16.4) | 68.6 (65.9-71.3) | 7.7 (6.4-9.3) |
| Gender |  |  |  |  |  |  |
| Cisgender Men (reference) | 18.1 (16.2-20.1) | 13.3 (11.5-15.2) | 35.9 (33.4-38.4) | 17.1 (15.2-19.2) | 53.2 (50.4-56.0) | 10.9 (9.3-12.7) |
| Cisgender Women | 16.2 (14.8-17.8) | 11.2 (9.9-12.6) | 35.9 (33.9-37.8) | 12.1 (10.9-13.4) | 56.1 (53.9-58.3) | 8.7 (7.6-10.1) |
| Transgender, Non-binary, or Other | 13.6 (6.6-25.9)¹ | 16.4 (8.8-28.4)¹ | 39.5 (27.2-53.3)¹ | 12.4 (7.0-20.9) | 50.4 (39.1-61.7)¹ | 12.6 (6.8-22.2)¹ |
| Education |  |  |  |  |  |  |
| Less Than HS | 14.7 (11.7-18.2) | 13.1 (10.3-16.7) | 27.3 (22.7-32.5) | 12.4 (9.4-16.2) | 39.1 (34.4-44.0) | 11.3 (8.7-14.7) |
| HS or Equivalent (reference) | 17.6 (15.0-20.5) | 11.6 (9.4-14.3) | 29.5 (26.3-33.0) | 13.2 (11.0-15.8) | 43.3 (39.7-47.0) | 10.4 (8.4-12.7) |
| Some College | 19.1 (16.6-21.9) | 10.8 (8.8-13.1) | 33.5 (30.4-36.7) | 14.6 (12.4-17.2) | 52.8 (49.0-56.7) | 11.6 (9.3-14.4) |
| College Graduate | 16.6 (15.0-18.4) | 13.1 (11.5-14.9) | 45.3 (43.2-47.4) | 16.0 (14.4-17.7) | 69.3 (66.9-71.6) | 7.6 (6.4-9.0) |
| Household Poverty |  |  |  |  |  |  |
| <100% | 19.8 (17.1-22.8) | 12.1 (9.9-14.8) | 27.4 (24.2-30.8) | 13.2 (10.9-15.8) | 40.0 (36.7-43.4) | 11.4 (9.4-13.8) |
| 100%-<200% (reference) | 14.3 (11.8-17.2) | 12.2 (9.9-15.0) | 31.4 (28.0-35.0) | 15.6 (13.2-18.3) | 47.3 (43.3-51.4) | 12.0 (9.8-14.7) |
| 200%-<400% | 17.5 (14.5-21.0) | 12.3 (9.2-16.3) | 40.3 (36.8-43.9) | 16.1 (13.5-19.1) | 52.4 (48.0-56.8) | 9.4 (7.2-12.1) |
| 400%-<600% | 15.9 (13.5-18.6) | 11.5 (9.3-14.0) | 36.6 (33.0-40.4) | 14.2 (11.4-17.5) | 71.6 (67.5-75.3) | 7.9 (5.8-10.8) |
| >= 600% | 17.1 (15.0-19.5) | 13.0 (11.1-15.2) | 47.1 (43.9-50.2) | 13.5 (11.6-15.6) | 71.8 (68.6-74.8) | 7.2 (5.6-9.1) |
| Notes: Data are from the NYC Community Health Survey 2022-2023. Data are weighted to the adult residential population per the American Community Survey. Confidence Intervals are a measure of estimate precision: the wider the CI, the more imprecise the estimate.  Prior COVID-19 ascertained by self-report: "From February 2020 until now, do you think you may have had COVID-19?"  ¹Estimate should be interpreted with caution. Estimate’s Relative Standard Error (a measure of estimate precision) is greater than 30%, or the 95% Confidence Interval half-width is greater than 10 or the sample size is too small, making the estimate potentially unreliable. | | | | | | |
